# Alzheimer’s disease and psychiatric polygenic risk are disease-specific and independent of plasma biomarkers in a real-world young-onset dementia cohort

**DOI:** 10.64898/2026.10.31.26366908

**Authors:** Oneil Girish Bhalala, Longfei Wang, Zachary F. Gerring, Jiru Han, Matthew JY Kang, Brett O. Trounson, Samantha M. Loi, Dennis Velakoulis, Dhamidhu Eratne, Melanie Bahlo, The MiND Study Group

## Abstract

**Background:** Young-onset dementia is frequently misdiagnosed, particularly where neurodegenerative and primary psychiatric disorders (PPD) overlap. Plasma biomarkers measure current pathology, whereas polygenic risk scores (PRS) capture lifelong genetic risk. Whether PRS derived from late-onset and population samples transfer to young-onset presentations, and how they relate to plasma biomarkers, is unknown.

**Methods:** We studied 687 participants of European ancestry from the Markers in Neuropsychiatric Disorders (MiND) Study, a prospective young-onset neuropsychiatry cohort: 195 healthy controls (HC), 283 PPD, 55 Alzheimer’s disease (AD) and 154 non-AD neurodegenerative disease (oNDG). PRS for AD, a cross-disorder psychiatric factor (PSY), frontotemporal dementia, Lewy body dementia and Parkinson’s disease were computed with PRS-CS, excluding the *APOE* locus. Plasma NfL, GFAP and p-tau217 were available for 332 participants. Models were adjusted for age, sex and *APOE* ε4 dose.

**Results:** Relative to HC, the AD PRS was associated with AD (OR 1.69 per SD, 95% CI 1.17–2.45) but not PPD (0.95, 0.77–1.16). The PSY PRS showed the reciprocal pattern (PPD 1.29, 1.05–1.58; AD 1.00, 0.72–1.40). No PRS was associated with any plasma protein. In distinguishing PPD from oNDG, NfL was the strongest discriminator (OR 2.15, 1.62–2.83). The AD PRS was also associated with oNDG (1.38, 1.11–1.71), without improving discrimination (AUC 0.764 for NfL alone versus 0.773 combined).

**Conclusion:** In young-onset presentations, AD and psychiatric polygenic risk are disease-specific and independent of plasma biomarkers of neurodegeneration. Their combined diagnostic value requires evaluation in larger, longitudinal pre-clinical cohorts.

## Introduction

Young-onset dementia, with symptom onset before 65 years, has an estimated global prevalence of 119 per 100,000.^1^ It arises from a heterogeneous group of neurodegenerative disorders, including Alzheimer’s disease (AD), whose early features often overlap with primary psychiatric disorders (PPD).^2^ Diagnosis in this age group is often incorrect, necessitating diagnostic revision after further observation and testing.^3^ Nearly one-third of individuals with early neurodegenerative disease receive a prior psychiatric diagnosis^4^, and nearly 40% of those presenting with young-onset neurocognitive symptoms have their initial diagnosis revised during follow-up.^5^ Time to diagnosis is longer than in late-onset dementia.^6^

Plasma biomarkers have begun to address this problem. Phosphorylated tau 217 (p-tau217) is highly specific for AD pathology and distinguishes AD from other neurodegenerative disorders, including in a young-onset clinical cohort.^7–9^ Neurofilament light chain (NfL), a non-specific marker of neuroaxonal injury, separates neurodegenerative disorders from PPD.^10–12^ Glial fibrillary acidic protein (GFAP), a marker of astrocytic activation and neuroinflammation, has shown less consistent discriminative value.^13^

In the Markers in Neuropsychiatric Disorders (MiND) Study, a real-world clinical cohort of young-onset neurocognitive symptoms^14^, plasma p-tau217 distinguished AD from behavioural-variant frontotemporal dementia (bvFTD) and from PPD with accuracies of 96% and 93%. NfL best separated all neurodegenerative disorders from PPD, while GFAP demonstrated minimal additional value.^15^ Nonetheless, nearly half of those with bvFTD had normal p-tau217 and NfL profiles. Protein biomarkers therefore leave a proportion of neurodegenerative presentations unresolved. As measures of current pathology, they also capture dynamic disease state rather than underlying susceptibility.^16^

Genetics offer a complementary, static measure of the baseline disease liability.^16^ Beyond *APOE*, the strongest risk factor for AD, successive genome-wide association studies (GWAS) of AD have expanded the number of associated risk loci.^17–19^ Polygenic risk scores (PRS) aggregate these effects into an individual measure of genetic liability. GWAS also exist for non-AD causes of dementia including FTD^20^, Lewy body dementia^21^, and Parkinson’s disease.^22^ These scores are derived predominantly from late-onset or population samples, and their transferability to young-onset clinical presentations is uncertain. Some studies support a shared common-variant architecture between late- and early-onset AD^23–25^, whereas others find little contribution of late-onset polygenic risk.^26,27^ For psychiatric illness, genomic structural equation modelling across 14 childhood- and adult-onset disorders has defined a common genetic factor shared across psychiatric diagnoses.^28^ The psychiatric common-factor PRS has not been examined in a dementia diagnostic setting.

How genetic liability relates to plasma protein markers is also unresolved. In older and population-based cohorts, the genetic contribution to plasma biomarker levels is carried largely by *APOE*. An *APOE*-excluded AD PRS showed no association with p-tau217, Aβ42/40, NfL or GFAP across ages 30–80^29^, and associations elsewhere have been confined to specific subgroups or outcomes.^30,31^ Combined genetic risk scores and plasma biomarkers appear to carry partly distinct information in late-onset dementia.^32,33^ Whether genetic and protein markers are independent in a young-onset population, and whether this extends to psychiatric liability, is unknown.

We aimed to address these gaps through analysis of a prospective cohort of patients referred to a tertiary young-onset neuropsychiatry service. In this study, we set out to test whether disease-specific PRS mark their corresponding clinical diagnoses and are independent of plasma NfL, GFAP and p-tau217. We also tested whether NfL and AD PRS contribute independently to distinguishing PPD from non-AD neurodegeneration, a common and challenging clinical scenario.

## Methods

### Participant Recruitment

Participants with young-onset neurocognitive and psychiatric symptoms referred to a tertiary center for diagnostic work-up were prospectively recruited for the MiND Study.^3,11,12,15^ After a follow-up period of over one year, a diagnosis was determined using consensus criteria, with details published prior.^15^

This study, part of the Markers in Neuropsychiatric Disorders Study (The MiND Study), was approved by the Melbourne Health Human Research Ethics Committee (2016.038, 2017.090, 2018.371, 2020.142) and the Walter and Eliza Hall Institute Human Research Ethics Committee (project ID 23/27).

### Blood Protein Biomarker Quantification

Blood protein biomarkers for NfL, GFAP, and p-tau217 were quantified as previously described.^15^ Briefly, EDTA plasma samples were collected during the diagnostic work-up and stored at −80C. Plasma NfL and GFAP were measured on a Quanterix Single molecule array (Simoa) HD-X analyzer. Plasma p-tau217 was measured using the University of Gothenburg assay.^34^

### Genotyping and Imputation

Genotyping and sequencing were performed by the Australian Genome Research Facility. 787 participants were genotyped on the Illumina Infinium Global Screening Array v3.0 with Multi-Disease and Psychiatric content add-ons, and a separate 141 participants underwent Illumina short-read whole-genome sequencing (approximately 100 Gbp per sample), joint-called with GATK.^35^

Array batches were quality-controlled individually in PLINK 2.0^36^, removing samples and variants with more than 10% missing genotypes (no samples were removed). Batches were then merged and the combined array set re-filtered for variant missingness (--geno 0.2) and Hardy–Weinberg equilibrium (--hwe 1e-5 0.001 midp; autosomal p < 1×10, X-chromosome female p < 1×10 ³, mid-p correction). The harmonised array-plus-sequencing dataset was restricted to array markers, merged, and then filtered for variant missingness (--geno 0.05), sample missingness (--mind 0.05), minor allele frequency (--maf 0.01), and Hardy–Weinberg equilibrium (--hwe 1e-6), yielding 928 samples and 563,455 variants submitted for imputation.

Phasing and imputation were performed on the TOPMed Imputation Server^37^ against the TOPMed r3 reference panel (GRCh38)^38^, with Eagle2 phasing^39^ and Minimac4 imputation (accessed January 2026). Imputed variants were retained under a frequency-dependent quality rule: directly typed variants at MAF > 5×10□□; imputed variants at MAF > 0.01 with R² > 0.5, or at 5×10□□ < MAF ≤ 0.01 with R² > 0.8. A final PLINK 2.0 pass (--geno 0.05, --maf 0.01, --hwe 1e-6) produced the analysis-ready set of 928 samples and 8,141,500 variants at MAF ≥ 1% across the MiND cohort.

Genetic ancestry was inferred in KING^40^ against the 1000 Genomes reference panel.^41^ For this study analyses were restricted to participants of inferred European ancestry in four pre-specified diagnostic groups totaling 687 participants: healthy controls (HC, n = 195), primary psychiatric disorder (PPD, n = 283), Alzheimer’s disease (AD, n = 55), and non-AD neurodegenerative disease (oNDG, n = 154). oNDG group consisted of frontotemporal dementia (FTD) n = 34, Huntington’s disease n = 24, Lewy body dementia (LBD) n = 10, Parkinson’s disease (PD) n = 4, and other dementia n = 82. Genotype source varied by diagnostic group: HC and PPD were array-genotyped, AD was sequenced, and oNDG comprised both (array n = 76, sequenced n = 78); this was pre-determined based on prior MiND Study projects.

*APOE* genotype was determined from two SNPs in the imputed dataset: rs429358 and rs7412. Alleles were assigned to each participant and the ε4 allele dose (0, 1, or 2) was used for downstream analyses.

### Polygenic Risk Score

Polygenic risk scores (PRS) were computed with PRS-CS, which applies a Bayesian continuous-shrinkage prior to SNP effect sizes using an external linkage-disequilibrium (LD) reference.^42^ Before scoring, the *APOE* locus (chr19:44–46 Mb, GRCh38) was excluded from GWAS summary-statistics files. For each trait, the effective sample size reported in the original GWAS was supplied as the PRS-CS N. Posterior SNP weights were estimated per chromosome (1–22) against the 1000 Genomes European LD reference, with the global shrinkage parameter learned from the data (PRS-CS auto). Per-chromosome posterior weights were concatenated and scores computed in the TOPMed-imputed, quality-controlled MiND participants genotypes with PLINK 2.0 using allele-matched effect sizes. PRS were calculated using summary statistics for AD^17–19^, PSY (cross-disorders P-factor)^28^, FTD^20^, LBD^21^, and PD.^22^

Each score was adjusted for residual population structure by regression on the first ten genetic principal components, and the residuals standardised to mean 0 and standard deviation (SD) 1 in healthy controls.

### Statistical Analysis

Analyses were performed in R 4.5.3 and were restricted to the European-ancestry subset. Tests were two-sided with a nominal significance threshold of 0.05. We distinguish nominal p-values (*p_nom_*) from Benjamini-Hochberg false-discovery-rate-adjusted p-values (*p_adj_*) throughout. False-discovery-rate correction was applied within pre-specified families: for each biomarker or polygenic risk score (PRS), the family comprised its three group contrasts against healthy controls (HC) within a given covariate set; for the primary psychiatric disorder (PPD) versus non-AD neurodegenerative disease (oNDG) univariate analysis, the family comprised the candidate predictors within a covariate set.

Each PRS was expressed in SD units of the European-ancestry HC distribution. Plasma biomarkers were natural-log-transformed and expressed as z-scores relative to the European-ancestry HC mean and SD; untransformed concentrations (pg/mL) were used for descriptive summaries and figures. Unless stated otherwise, models were adjusted for age at blood draw, sex, and *APOE* ε4 allele dose.

#### Cohort description

Descriptive statistics are medians (interquartile range) for continuous variables and counts (percentages) for categorical variables. Unadjusted differences across the four diagnostic groups were tested by the Kruskal-Wallis test for continuous variables (age) and the Pearson chi-square test for categorical variables (sex, *APOE* ε4 carriership). Availability of the plasma panel by diagnostic group was compared by chi-square test.

#### Polygenic risk scores

Five candidate AD PRS^17–19^ builds were compared for their discrimination of AD from HC by logistic regression (odds ratio [OR] per SD, adjusted for age, sex, and *APOE* ε4 dose, with Wald 95% confidence intervals [CIs]). The best-separating build (2026 Consensus No-Biobanks)^19^ was carried forward for all subsequent AD PRS analyses.

Mean differences of each disease PRS from HC in each diagnostic group were estimated by linear regression of the standardised PRS on diagnostic group with covariates, using ordinary least squares.

Condition specificity of the AD and psychiatric (PSY)^28^ genetic-liability axes was assessed in four separate two-group logistic models, pairing each score (AD PRS, PSY PRS) with each contrast (AD versus HC, PPD versus HC), each fitted on its own complete-case sample and adjusted for covariates. Each model consisted of the OR per SD (Wald 95% CI), the apparent area under the receiver operating characteristic curve (AUC) of the PRS-plus-covariate model with DeLong 95% CIs, and the apparent AUC of a covariate-only model.

#### Plasma biomarkers

Plasma NfL, GFAP, and p-tau217 concentrations were summarised by diagnostic group as medians (interquartile range) on the untransformed (pg/mL) scale.

#### Protein and genetic biomarker interactions

Pairwise associations among the three plasma proteins and five disease PRS were assessed by partial Spearman correlation, with Benjamini-Hochberg correction applied within each matrix.

#### Discrimination of PPD from oNDG

Two complete-case cohorts were defined. In the maximum-n cohort, each predictor was modelled in the largest sample that supported it (genotype for PRS, the relevant assay for protein biomarkers). The primary cohort was complete on the pre-specified predictors (NfL and AD PRS) and covariates; the joint model and all discrimination statistics were fitted to this cohort.

Each predictor was first tested univariately by logistic regression (OR per SD, adjusted for covariates), in both cohorts. The joint multivariable model entered NfL and AD PRS simultaneously together with covariates, allowing each marker to be assessed adjusted for the other. Discrimination was summarised by the AUC of four nested models (covariates only; NfL; AD PRS; NfL plus AD PRS): apparent AUCs are shown, and both apparent and optimism-corrected AUCs (Harrell bootstrap, 1000 resamples, non-converged resamples excluded) were calculated.

## Results

### Description of the MiND Genomics Study Demographics

The analysis cohort comprised 687 participants of European ancestry: 195 healthy controls (HC), 283 primary psychiatric disorder (PPD), 55 Alzheimer’s disease (AD), and 154 non-AD neurodegenerative disease (oNDG) (Table 1). The groups differed in age at blood draw (median 58.9 [IQR: 46.7 - 66.5] years overall, with PPD youngest at 53.5 [IQR: 40.5 - 62.6] and HC oldest at 62.0 [IQR: 51.4 - 69.0]; *p_nom_* < 0.001), sex (60% female overall, ranging from 45% in oNDG to 78% in HC; *p_nom_* < 0.001), and *APOE* ε4 dose (66% non-ε4 carrier overall, lowest in AD at 44%; *p_nom_* < 0.001) (Table 1). A complete plasma panel (NfL, GFAP, p-tau217) was available for 332 participants (48%), while NfL and GFAP were available for 371 participants (54%).

**Table 1.** MiND Genomics Study Demographics.

|  | **Overall<br>N = 687** | **HC<br>N = 195** | **PPD<br>N = 283** | **AD<br>N = 55** | **oNDG<br>N = 154** |
|---|---|---|---|---|---|
| **Age at blood draw<sup>1</sup>** | 58.9 (46.7, 66.5) | 62.0 (51.4, 69.0) | 53.5 (40.5, 62.6) | 61.8 (58.6, 67.7) | 60.5 (47.6, 67.1) |
| **Female** | 414 (60%) | 153 (78%) | 164 (58%) | 27 (49%) | 70 (45%) |
| ***APOE* ε4 Dose** |  |  |  |  |  |
| **0** | 454 (66%) | 132 (68%) | 201 (71%) | 24 (44%) | 97 (63%) |
| **1** | 211 (31%) | 61 (31%) | 76 (27%) | 23 (42%) | 51 (33%) |
| **2** | 22 (3.2%) | 2 (1.0%) | 6 (2.1%) | 8 (15%) | 6 (3.9%) |
| **Protein biomarker availability** |  |  |  |  |  |
| **NfL/GFAP/P-tau217** | 332 (48%) | 118 (61%) | 83 (29%) | 30 (55%) | 101 (66%) |
| **NfL/GFAP** | 371 (54%) | 124 (64%) | 97 (34%) | 35 (64%) | 115 (75%) |

<sup>1</sup> *Median (interquartile range)*
*HC, healthy control; PPD, primary psychiatric disorder; AD, Alzheimer's disease; oNDG, non-AD neurodegenerative disease.*

### Polygenic Risk Scores Demonstrate Disease Specificity

We first compared five recent AD PRS builds^17–19^ for their ability to separate AD from HC within the MiND genomics study (per SD increment, adjusted for age, sex, and APOE; Supplementary Figure 1, Supplementary Table 1). The Kunkle 2019^17^ build did not discriminate (OR 1.18 per SD [95% CI: 0.84 - 1.66], *p_nom_* = 0.33), whereas the 2026 Consensus builds^19^ were the most robust within the cohort, with the No-Biobanks build yielding the strongest separation (OR 1.69 [95% CI: 1.17 - 2.45], *p_nom_* = 0.005). The No-Biobanks Consensus build was carried forward for all subsequent AD PRS analyses.

Across the four diagnostic groups, certain disease PRS showed condition-specific discrimination when compared to HC (Figure 1, Supplementary Figure 2, Supplementary Tables 2 and 3 [*APOE* ε4 adjusted]). AD PRS^19^ was highest for AD (mean +0.49 SD relative to HC, *p_adj_* < 0.001) and intermediate for oNDG (+0.23 SD), while PPD was not elevated compared to HC (−0.05 SD).

**Figure 1.**
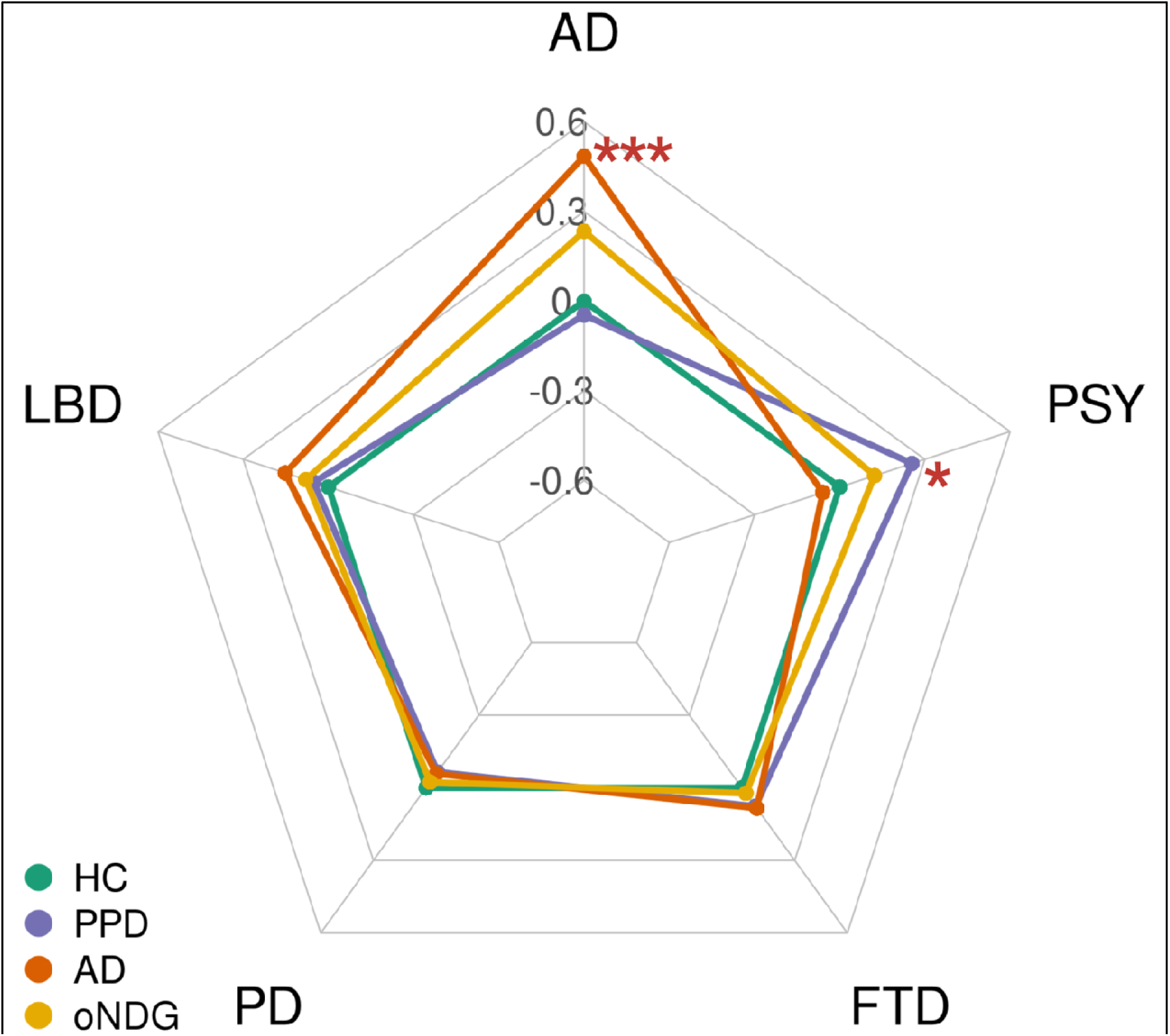
Radar plot of disease-specific PRS. Mean polygenic risk score (standardised to HC, adjusted for age and sex) on each disease PRS axis (AD, PSY, FTD, PD, LBD) for the four diagnostic groups (coloured lines). *** *p_adj_* < 0.001, * *p_adj_* < 0.05. HC, healthy control; PPD, primary psychiatric disorder; AD, Alzheimer’s disease; oNDG, non-AD neurodegenerative disease; PSY, psychiatric; FTD, frontotemporal dementia; PD, Parkinson’s disease; LBD; Lewy body dementia.

PSY PRS^28^ showed the reciprocal pattern: highest in PPD (+0.25 SD, *p_adj_* < 0.05) and at the HC level in AD (0.02 SD). The FTD^20^, PD^22^, and LBD^21^ PRS were comparatively equivalent to HC across the diagnostic groups.

In comparing AD and PPD diagnostic categories, AD PRS and PSY PRS each discriminated only in its own diagnostic contrast (Figure 2, Supplementary Table 4). AD PRS discriminated AD from HC (OR 1.69 per SD [95% CI: 1.17 - 2.45], *p_nom_* = 0.005) but not PPD from HC (OR 0.95 [95% CI: 0.77 - 1.16], *p_nom_* = 0.60), whereas PSY PRS discriminated PPD from HC (OR 1.29 [95% CI: 1.05 - 1.58], *p_nom_* = 0.013) but not AD from HC (OR 1.00 [95% CI: 0.72 - 1.40], *p_nom_* = 0.99); neither PRS was associated with the other diagnostic group. Similarly, relative to a model with age, sex, and *APOE* ε4 dose alone, each PRS improved the AUC only for its own contrast and by a small margin (AD PRS in AD, 0.73 to 0.76; PSY PRS in PPD, 0.70 to 0.71), while the contrasting-comparison AUCs were unchanged (0.73 and 0.70). The genetic associations were therefore condition-specific but added little discrimination beyond demographic covariates.

**Figure 2.**
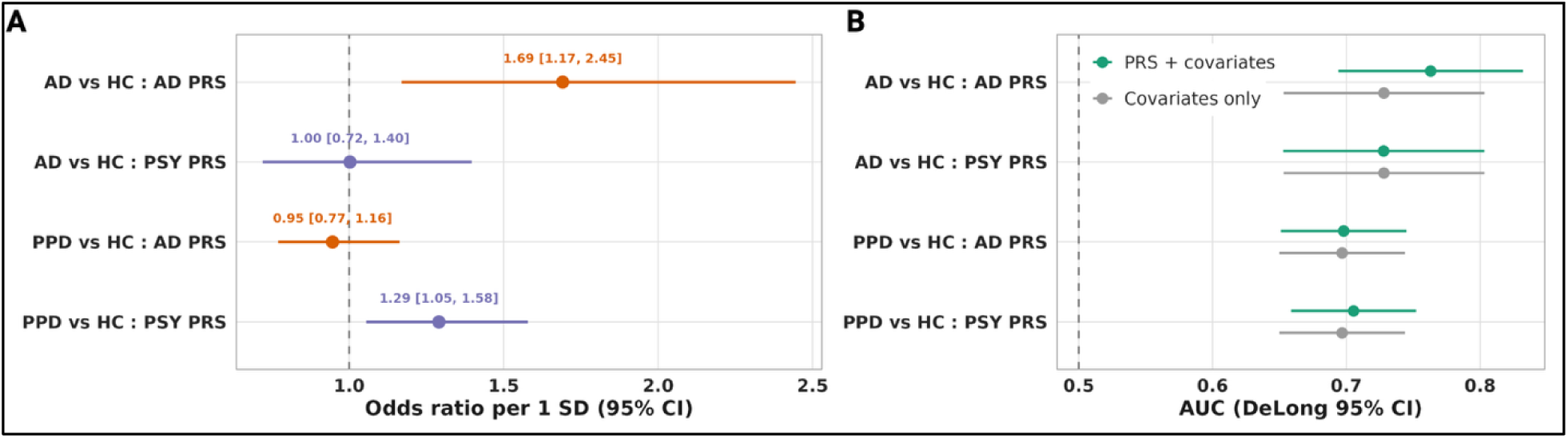
Condition-specific associations of the AD and psychiatric genetic-liability axes. Each row is a diagnostic contrast paired with one polygenic risk score, adjusted for age, sex, and *APOE* ε4 dose. (A) Odds ratio per 1 SD of PRS [95% CI]; the dashed line marks the null (OR = 1). (B) Discrimination for the PRS + covariate model (green) versus the covariate-only model (grey), AUC with DeLong 95% CI. HC, healthy control; PPD, primary psychiatric disorder; AD, Alzheimer’s disease.

### Protein and Genetic Biomarkers are Independent

Plasma biomarker distributions followed the expected neurodegeneration pattern and resembled those from the larger MiND clinical cohort (Supplementary Figure 3, Supplementary Tables 5 and 6).^15^ All three markers were highest in AD (median NfL 21.5 pg/mL, GFAP 244 pg/mL, p-tau217 3.33 pg/mL). NfL was higher in neurodegenerative disorders (AD 21.5, oNDG 19.4, HC 13.0, PPD 10.5 pg/mL). P-tau217 demonstrated AD-specificity (3.33 pg/mL in AD; ranging between 0.85 and 1.09 pg/mL in the other three groups). PPD had the lowest NfL levels of any group.

We next examined whether the genetic and protein biomarkers were correlated. After adjusting for age, sex, *APOE* ε4 dose, and diagnosis, the only associations to survive correction were among the three protein markers (NfL–GFAP ρ = 0.40, GFAP–p-tau217 ρ = 0.32, NfL–p-tau217 ρ = 0.25; all *p_adj_*< 10□□), with one weak correlation between AD PRS and FTD PRS (ρ = 0.10, *p_adj_* = 0.043) (Supplementary Figures 4 and 5, Supplementary Table 7). No PRS was associated with any protein marker, whether estimated by partial rank correlation (all *p_adj_* > 0.4) or by fully adjusted linear regression, in which every PRS-to-protein coefficient was near zero (for example, AD PRS with NfL β = −0.005, *p_adj_*= 0.99; all *p_adj_* > 0.8) (Supplementary Table 8). Within this cohort, the disease genetic liability captured by the PRS and the protein biomarker levels were statistically independent.

### NfL and AD PRS Independently Discriminate Psychiatric Disorders and non-AD Neurodegeneration

A diagnostic challenge in young-onset presentations is separating PPD from non-AD neurodegeneration, where symptoms often overlap. We used this clinical dilemma as motivation to determine the contributions of protein and genetic biomarkers in differentiating between these two clinically-ambiguous conditions.

Univariate and multivariable analyses demonstrated that both NfL levels and AD PRS were independent discriminators of PPD from oNDG. NfL provided the strongest separation with univariate analysis (OR 2.15 per SD [1.62 - 2.83], *p_nom_* = 7.5 × 10□□, *p_adj_* = 6.0 × 10□^7^; adjusted for age, sex, and *APOE* ε4 dose; Figure 3A, Supplementary Table 9). AD PRS was nominally associated with discrimination but did not survive correction (OR 1.38 [1.03 - 1.85], *p_nom_* = 0.032; *p_adj_* = 0.13), likely due to the smaller sample size (n = 97 PPD, 115 oNDG) with both protein and genetic biomarkers. Note that in the full genotyped cohort (n = 283 PPD, 154 oNDG), AD PRS was significantly discriminatory between PPD and oNDG (OR 1.38 per SD [1.11 - 1.71], *p_adj_* = 0.013; Supplementary Table 10). No other protein markers or disease PRS discriminated the two groups (all *p_nom_* and *p_adj_* were > 0.05).

**Figure 3.**
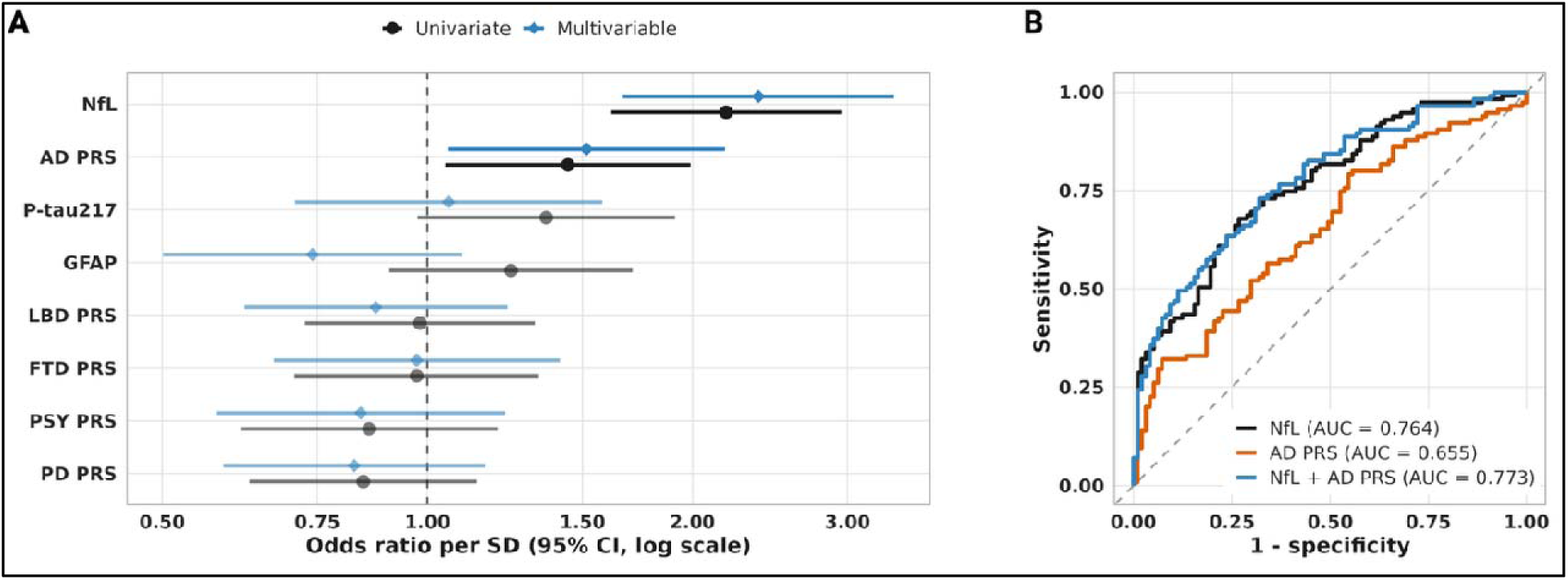
NfL and AD PRS as discriminators of PPD from oNDG. (A) Odds ratio per 1 SD for each protein and genetic predictor discriminating oNDG from PPD, adjusted for age, sex, and *APOE* ε4 dose (log scale; dashed line, OR = 1). Univariate estimates (black) are each predictor modelled alone; multivariable estimates (blue) are from a single model containing all predictors simultaneously. (B) Receiver operating characteristic curves (AUC) for NfL, AD PRS, and NfL + AD PRS. HC, healthy control; PPD, primary psychiatric disorder; oNDG, other neurodegenerative disease; NfL, neurofilament light; AD PRS, Alzheimer’s disease polygenic risk score.

In the joint multivariable model, both markers remained nominally significant and essentially unchanged when adjusting for each other (NfL OR 2.14 per SD [1.62 - 2.83], *p_nom_*= 1.0 × 10□□; AD PRS OR 1.40 [1.01 - 1.93], *p_nom_*= 0.044; Figure 3A), consistent with the two variables being independent of each other (Supplementary Figure 4). Because the two variables are orthogonal and NfL is much the stronger, adding AD PRS did not improve discrimination over NfL alone (AUC 0.764 for NfL, 0.773 for NfL + AD PRS, 0.655 for AD PRS alone; Figure 3B, Supplementary Table 11).

## Discussion

We examined disease-specific genetic liability alongside plasma NfL, GFAP and p-tau217 in a real-world young-onset dementia cohort and observed three main findings. First, genetic liability to AD and to PPD were distinct. The AD PRS^19^ was associated with AD but not PPD relative to HC, and the psychiatric common-factor PRS^28^ (which represents the shared genetic architecture amongst psychiatric diseases) showed the reciprocal pattern. Second, genetic liability was statistically independent of plasma protein levels as the only significant correlations were among the proteins themselves. Third, in separating PPD from non-AD neurodegeneration, NfL was the dominant discriminator, and the AD PRS carried a small, but independent association. Together, these results indicate that genetic liability and plasma protein markers capture separable dimensions of disease, even in a diagnostically heterogeneous young-onset population where misdiagnosis and diagnostic delay are most common.

The most recent AD GWAS^19^ transferred well to the young-onset cohort studied here. AD PRS discrimination of AD from HC strengthened with each successive GWAS, from Kunkle 2019 (OR 1.18 per SD) to Bellenguez 2022 (OR 1.42) to the 2026 Consensus meta-analysis (OR 1.51–1.69). The three Consensus builds differ in their inclusion of proxy and biobank-derived cases, which has been shown to impact AD heritability estimates^43^, yet gave similar estimates in our study, likely reflecting insufficient power to distinguish effects of different case inclusions. The Bellenguez-derived estimate is of comparable magnitude to the 1.42 per SD reported for an 83-variant Bellenguez-derived score across European-ancestry cohorts, including 1.42 (1.26–1.59) in an Australian cohort.^44^ This suggests that PRS performance in MiND sits within the published range, although the two scores differ in construction (genome-wide PRS-CS versus lead variants). These findings are consistent with a shared common-variant architecture between late-onset and young-onset AD^23^, and with studies showing that late-onset AD PRS carry signal in younger cases.^24,25,45^ Transferability has not been uniform, however, with some studies finding little contribution of late-onset polygenic risk to early-onset or atypical AD.^26,27,46^ Further work, with increased cohort size, is needed to more precisely understand the genetic contribution of late-onset AD genetic liability in young-onset cases.

The association of the psychiatric common-factor PRS with PPD is, to our knowledge, the first demonstration of psychiatric genetic risk in a young-onset dementia diagnostic setting. The PSY PRS, derived from genomic structural equation modelling across 14 psychiatric disorders^28^, was associated with PPD relative to HC (OR 1.29 per SD) and showed no association with AD (OR 1.00). These findings suggest that a cross-disorder factor is applicable to a clinical PPD group that spans depressive, bipolar, psychotic and functional presentations. The reciprocal pattern is informative in both directions. Patients whose presentations were ultimately attributed to a psychiatric cause carried the polygenic signature of psychiatric illness. They did not carry excess AD liability (−0.05 SD relative to HC), in keeping with their low p-tau217 levels. The PSY PRS did not significantly discriminate between PPD and oNDG, a common clinical challenge (OR 0.88 [95% CI 0.71 – 1.09], Supplementary Table 10 for the full genotyped cohort). oNDG was intermediate between HC and PPD on the PSY PRS axis (+0.14 SD relative to HC), so the two groups differed by only 0.11 SD. The oNDG group also reflects the referral pattern of a tertiary neuropsychiatry service, with substantial representation of Huntington’s disease and heterogeneous non-AD dementias, and its psychiatric genetic liability may not generalise to neurodegenerative cohorts of different composition. Larger studies with adequately sized, diagnostically defined neurodegenerative groups are needed to establish whether psychiatric polygenic liability contributes to this distinction. Behavioural-variant FTD should be a priority, as a frequent mimic of primary psychiatric disorders, especially when NfL plasma levels may be in the normal range.^15^

The disease-PRS we tested were not associated with plasma protein levels. This aligns with recent work in older populations, in which the genetic contribution to plasma biomarker levels is carried largely by *APOE*. In 5,913 Netherlands Twin Register participants aged 30–80, an AD PRS (based on the Bellenguez GWAS) excluding *APOE* showed no association with p-tau217, Aβ42/40, NfL or GFAP concentrations across age, whereas ε4 carriage was associated with steeper age-related changes in Aβ42/40, p-tau217 and GFAP.^29^ Where associations with *APOE*-excluded scores have been reported, they have been confined to specific subgroups or outcomes. In ADNI, such an association with plasma p-tau181 was seen only in mild cognitive impairment and amyloid-positive participants, whereas the *APOE*-inclusive score was associated irrespective of diagnosis or amyloid status.^30^ In BIOCARD, the *APOE*-excluded score was related to longitudinal Aβ42/40 decline only among ε4 non-carriers.^31^ Conceptually, the PRS captures the fixed lifelong genetic liability, whereas plasma proteins reflect current neuropathology, so independence between the two is expected. Independence also implies that genetic and protein measures carry complementary information. Utility of these orthogonal signals is not clearly demonstrated in our or other’s work; how these signals can be exploited to obtain earlier and more accurate diagnoses, especially in prodromal or preclinical states remains to be determined.

The comparison of PPD with non-AD neurodegeneration was used to test independence directly, in the clinical contrast where symptoms most often overlap. NfL separated the groups strongly (OR 2.15 per SD; AUC 0.76), concordant with the plasma and CSF NfL literature from this service and the wider MiND clinical cohort^12,15^, which overlaps with the present sample. The AD PRS was also associated with oNDG relative to PPD (OR 1.38 per SD in the full genotyped cohort, Supplementary Table 10). It retained this association when modelled jointly with NfL, while discrimination changed little (AUC 0.764 for NfL, 0.773 combined). With orthogonal predictors and a much stronger protein marker, this non-additivity is not surprising. It confirms that the AD PRS carries information distinct from NfL that does not improve discrimination in clinically established cases. This supports further research into the utility of PRS prior to clinical diagnoses in a real-world clinical cohort.

This study has several limitations. Genotyping platform was aligned with diagnosis as AD participants underwent WGS, HC and PPD participants were array-genotyped, and oNDG participants were split across platforms. We mitigated this by restricting sequencing data to array sites before joint imputation. The null PSY, FTD, LBD and PD PRS contrasts between AD and HC also act as internal negative controls. Residual platform effects nevertheless cannot be fully excluded. The oNDG group reflects the referral pattern of the Royal Melbourne Hospital Neuropsychiatry Centre. It over-represents Huntington’s disease, with few FTD, LBD and PD cases, which limits power for the corresponding disease-specific PRS and generalisability to general memory clinics. The association of the AD PRS with oNDG may partly reflect AD co-pathology or unrecognised AD within oNDG, particularly as diagnoses were clinical and not all participants had CSF or PET confirmation. The AD PRS build was selected on the AD-versus-HC contrast, so the reported OR is likely optimistic, although all three Consensus builds gave overlapping estimates. The plasma panel was available for 48% of the cohort and less often in PPD (29%), so the primary PPD-versus-oNDG cohort was a selected subset. The AD PRS estimate was nevertheless the same in this subset and in the full genotyped cohort (OR 1.38 per SD in both). The AD group was modest (n = 55), and the design was cross-sectional so biomarker levels may be impacted by dementia severity in addition to the underlying pathology.

Analyses were restricted to European ancestry, thereby limiting PRS effect generalizability to other ancestral groups.

In summary, in a young-onset neuropsychiatry cohort, AD and psychiatric genetic liability each marked its own condition relative to healthy controls, and both were independent of plasma biomarkers of neurodegeneration. Genetic liability and protein biomarkers capture distinct dimensions of disease. Establishing whether this complementary structure translates into diagnostic benefit will require external validation, longitudinal follow-up with prodromal cases, and multi-ancestry cohorts.

## Data Availability

All data produced in the present study are available upon reasonable request to the authors

