## Supplemental Figures for "Alzheimer’s disease and psychiatric polygenic risk are disease-specific and independent of plasma biomarkers in a real-world young-onset dementia cohort"

**Supplementary Figures**

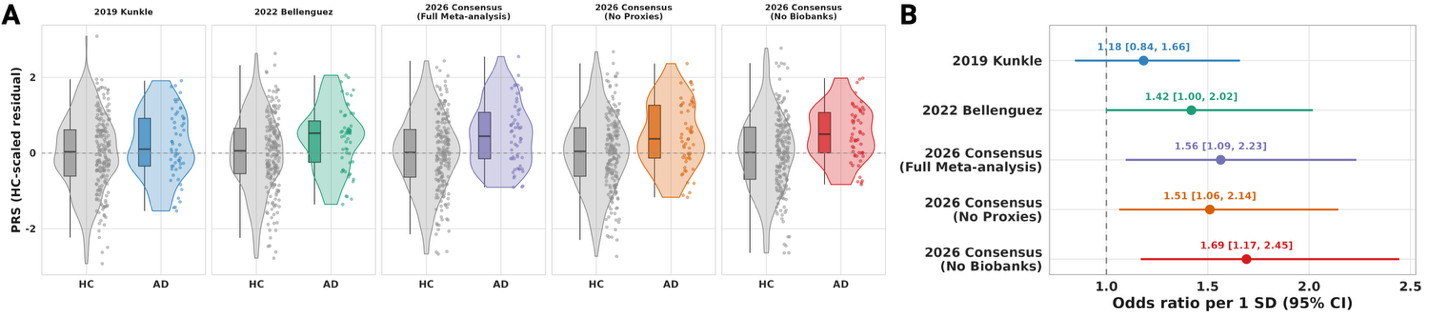

**Supplementary Figure 1 | Performance of recent AD PRS in MiND Cohort**. Five candidate AD PRS builds, all with the APOE locus excluded by chromosomal position, residualised and HC-scaled. (A) Distributions for HC and AD for each build (box, violin, and scatter points; dashed line = HC mean). (B) Discrimination of AD from HC, odds ratio per 1 SD [95% CI], adjusted for age, sex, and *APOE* ε4 dose.

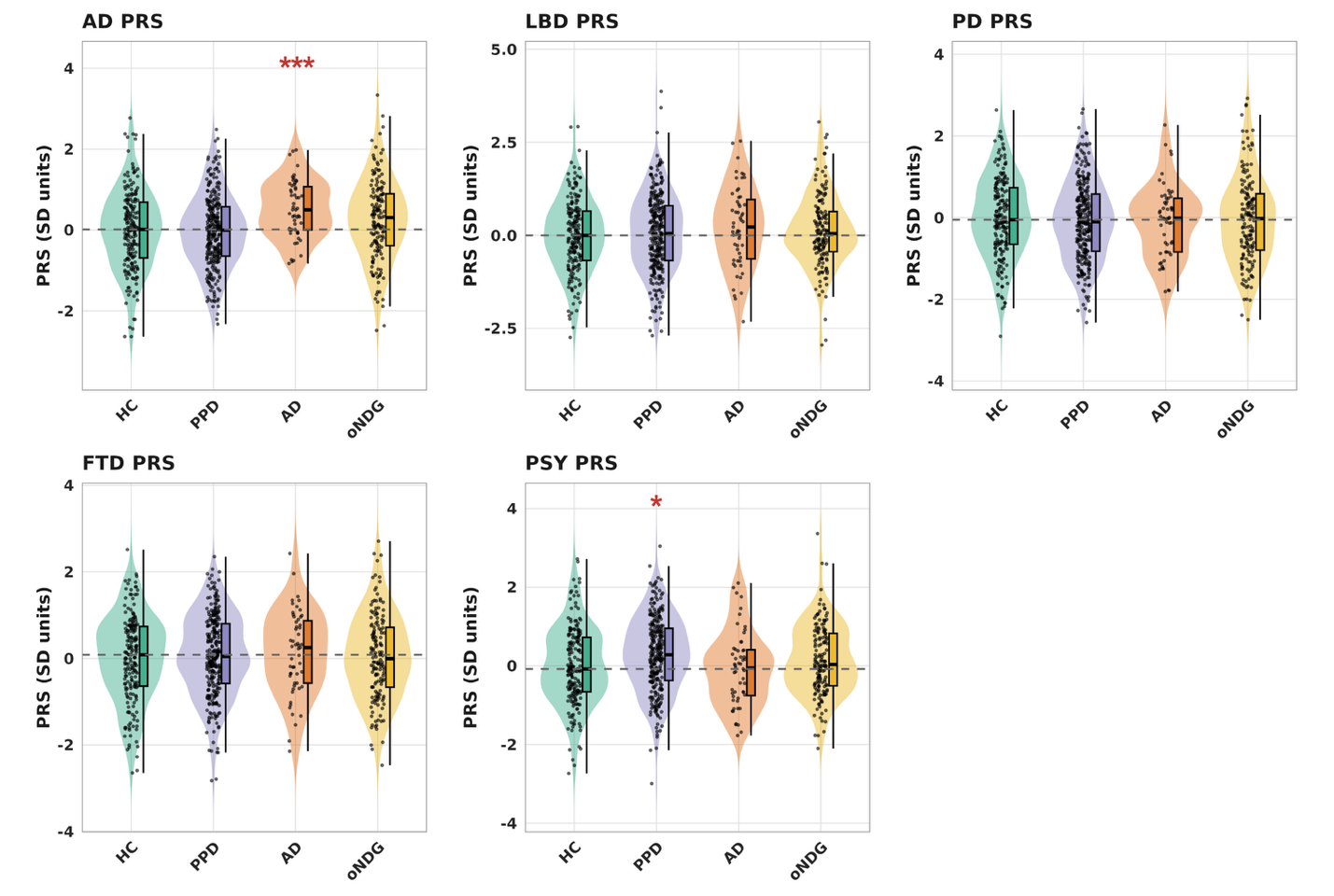

**Supplementary Figure 2 | Distribution of recent disease PRS across diagnostic categories**. Distribution of scaled disease-specific PRS, adjusted for age, sex, and *APOE* ε4 dose, across four diagnostic categories. AD PRS indicates the 2026 Consensus No-Biobanks PRS. Dashed line, HC median line. *** *p_adj_* < 0.001, * *p_adj_* < 0.05, compared to HC.

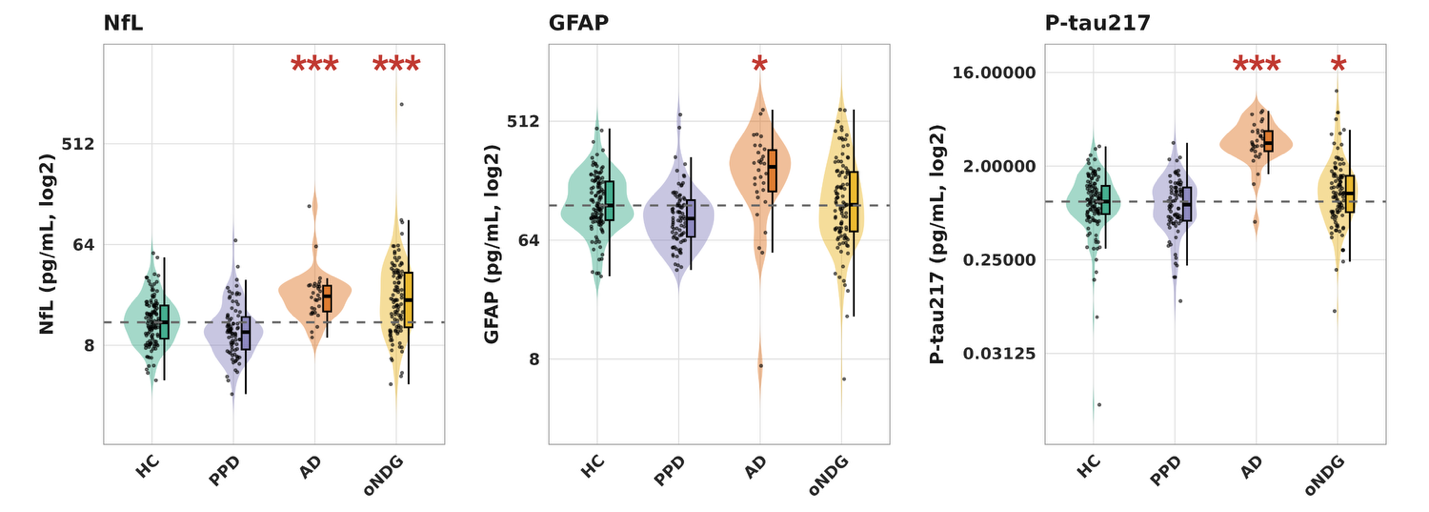

**Supplementary Figure 3 | Distribution of protein biomarkers across diagnostic categories**. Distribution of protein biomarkers, adjusted for age, sex, and *APOE* ε4 dose, across four diagnostic categories. Dashed line, HC median line ; log2 scale applied. *** *p_adj_* < 0.001, * *p_adj_* < 0.05, compared to HC.

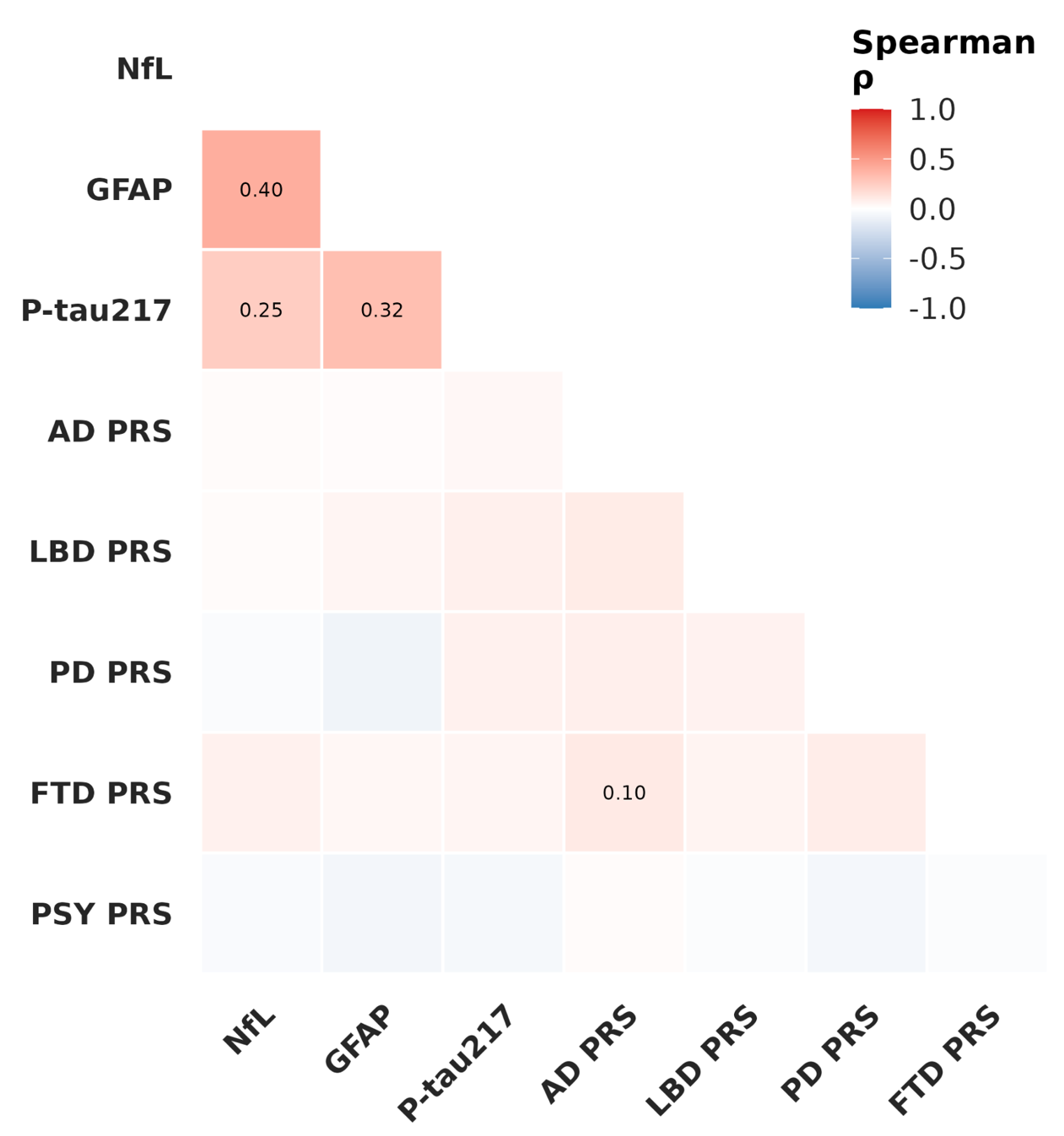

**Supplementary Figure 4 | Correlation between protein and genetic biomarkers**. Spearman r values, coloured by direction of association, between each pairwise comparison, adjusted for age, sex, *APOE* e4 dose and diagnoses. Only values with *P_adj_* < 0.05 are shown.

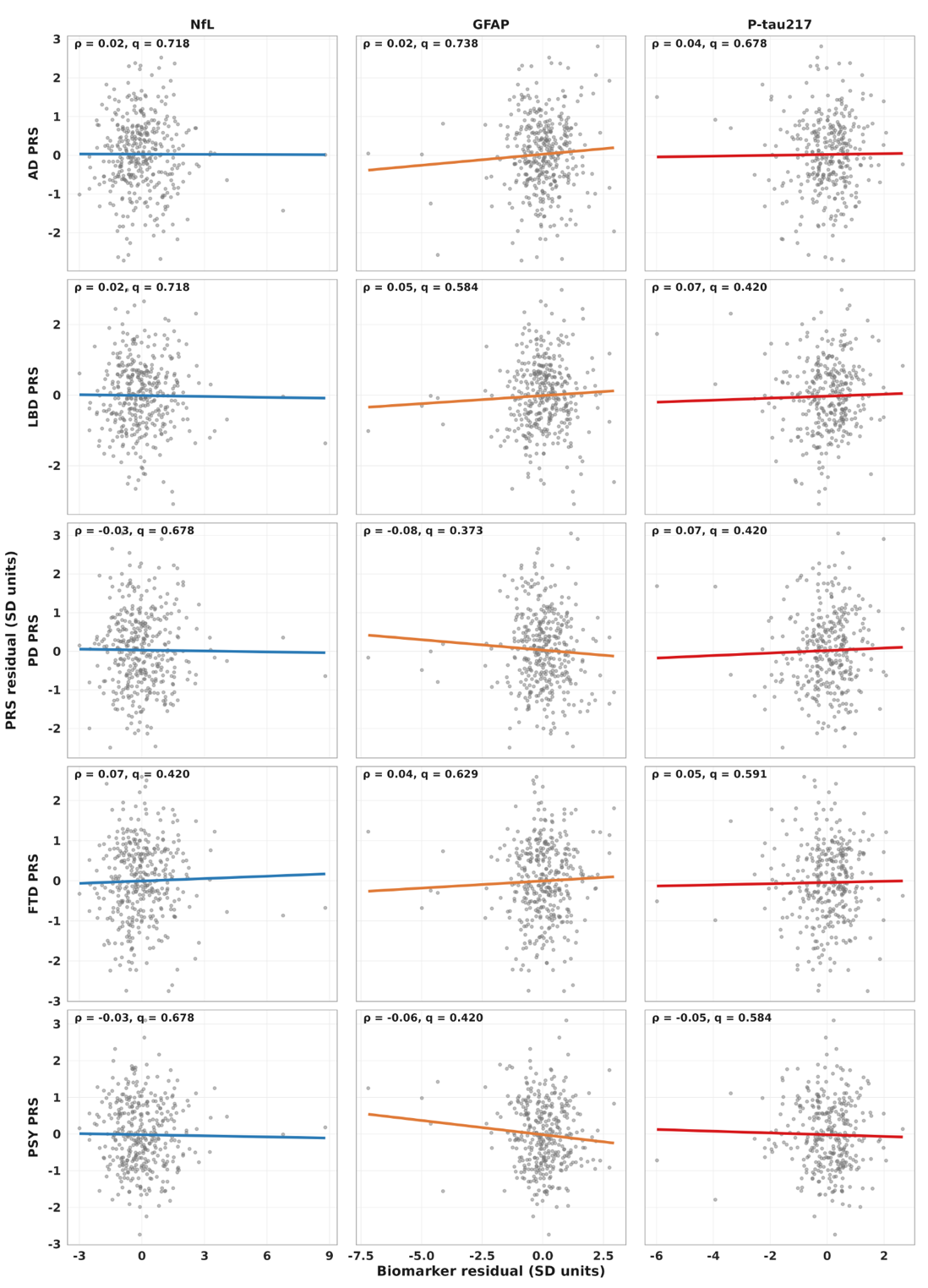

**Supplementary Figure 5 | Spearman correlation plots between protein and genetic biomarkers**. Fully adjusted (age, sex, *APOE* e4 dose, and diagnosis) Spearman correlation plots for each protein biomarker and disease PRS. r, Spearman coefficient; q, adjusted p-value. Line represents the simple linear regression for illustrative purposes.
